# Serious underlying medical conditions and COVID-19 vaccine hesitancy

**DOI:** 10.1101/2022.04.06.22273080

**Authors:** Daphne Day, Lisa Grech, Mike Nguyen, Nathan Bain, Alastair Kwok, Sam Harris, Hieu Chau, Bryan Chan, Richard Blennerhassett, Louise Nott, Nada Hamad, Annette Tognela, David Hoffman, Amelia McCartney, Kate Webber, Jennifer Wong, Craig Underhill, Brett Sillars, Antony Winkel, Mark Savage, Bao Sheng Loe, Daniel Freeman, Eva Segelov, the CANVACCS, DIABVACCS and MSVACCS investigators

## Abstract

**Objective:** To examine vaccine uptake, hesitancy and explanatory factors amongst people with serious and/or chronic health conditions, including the impact of underlying disease on attitudes to vaccination.

**Design:** Cross-sectional survey.

**Setting:** Ten Australian health services.

**Participants:** 4683 patients (3560 cancer, 842 diabetes and 281 multiple sclerosis) receiving care at the health services participated in the 42-item survey, between June 30 to October 5, 2021.

**Main outcome measures:** Sociodemographic and disease-related characteristics, COVID-19 vaccine uptake, and the scores of three validated scales which measured vaccine hesitancy and vaccine-related beliefs generally and specific to the participants’ disease, including the Oxford COVID-19 Vaccine Hesitancy Scale, the Oxford COVID-19 Vaccine Confidence and Complacency Scale and the Disease Influenced Vaccine Acceptance Scale. Multivariable logistic regression was used to determine the associations between scale scores and vaccine uptake.

**Results:** Of all participants, 81.5% reported having at least one COVID-19 vaccine. Unvaccinated status was associated with younger age, female sex, lower education and income, English as a second language, and residence in regional areas (all p<0.05). Unvaccinated participants were more likely to report greater vaccine hesitancy and more negative perceptions toward vaccines (all p<0.05). Disease-related vaccine concerns were associated with unvaccinated status and hesitancy, including greater complacency about COVID-19 infection, and concerns relating to vaccine efficacy and impact on their disease and/or treatment (all p<0.05).

**Conclusions:** Disease-specific concerns impact COVID-19 vaccine-related behaviours and beliefs in people with serious and/or chronic health conditions. This highlights the need to develop targeted strategies and education about COVID-19 vaccination to support medically vulnerable populations and health professionals.

**Trial registration:** ACTRN12621001467820

## Introduction

More than one in three people aged 16-years and over have at least one chronic health condition.^1^ This ‘medically vulnerable’ population has been disproportionately affected by the COVID-19 pandemic, with a higher risk of severe complications and death, as well as disruptions in usual care.^2^ Once available, vaccination has become one of the most critical public health defences in limiting the health and social impacts of the COVID-19 pandemic and to prevent infection and its significant sequelae, people with serious comorbidities have been prioritised in vaccination programs in many countries.^3^ Despite this, many countries reported slow uptake and a significant proportion of vaccine refusal, including in vulnerable populations.^4-6^ Reluctance to accept vaccines for disease prevention is not a new phenomenon. Vaccine hesitancy, defined as ‘delay in acceptance or refusal of vaccination despite availability’,^7^ was identified as one of the top ten threats to global health by the World Health Organisation in 2019.^8^

In the current pandemic, COVID-19 vaccine hesitancy rates have been reported in up to 50% of the general population, with significant regional variability.^4 5 9 10^ Vaccine hesitancy literature predating COVID-19 lists common associated factors including younger age, lower education and income levels, prior anti-vaccine views and low trust in governments.^4 5 9 10^ Public confidence in vaccine safety and efficacy has been particularly prominent in COVID-19 vaccine acceptance in the setting of rapid vaccine development, extensive media coverage of vaccine-related adverse events and proliferation of misinformation in the era of social media.^11^ Immunocompromised patients were underrepresented in the early COVID-19 vaccine registration trials,^12-14^ with vaccine efficacy and safety data in these cohorts emerging later.^15-17^ The uncertainty around vaccination for patients with serious underlying disease is reflected by disease-related concerns reported in several small survey-based studies^6 18 19^ and may be further compounded by the lack of tailored public health advice or inconsistent responses from clinicians. Considering the likely need for further vaccine doses (even regular boosters) due to the emergence of new variants and waning of existing immunity, there is an immediate and ongoing challenge for health care providers globally to understand drivers of vaccine hesitancy. This can be used to define the education needs in vulnerable populations to promote, maintain and in some cases, reinvigorate vaccination acceptance in this group.

Previously published COVID-19 vaccine hesitancy scales were developed in the general population prior to vaccine approval and widespread use.^9^ Our study took place as vaccines were instituted via nation-wide campaigns, and comprehensively assessed multiple measures of hesitancy and complacency, including the six-item Disease Influenced Vaccine Acceptance Scale (DIVAS-6) that we developed and validated. This comprises two distinct and addressable factors: perceived vulnerability to COVID-19 infection due to underlying chronic disease, and concerns regarding vaccine efficacy, safety and impact on disease control (manuscript under review). The aim of this large, multi-site Australian study undertaken in 2021 in people with cancer, diabetes and multiple sclerosis (MS) was to evaluate COVID-19 vaccine uptake, intent and hesitancy on the background of understanding general and disease-related beliefs regarding vaccine importance, benefit and safety.

## Methods

### Study design

This cross-sectional study was implemented at ten health services across four states in Australia, encompassing five metropolitan and five regional areas, across six public and four private settings, with a collective catchment population of 4.9 million. The survey opened at the coordinating site on June 30 2021 and concluded at all sites on October 5 2021. During this period, there were varying lockdowns, public health restrictions and vaccine rollout recommendations (Figure 1). The study was approved by the Monash Health Human Research Ethics Committee (RES-21-0000-364L – 76466) and registered with the Australian New Zealand Clinical Trials Registry (ACTRN12621001467820).

**Figure 1.**
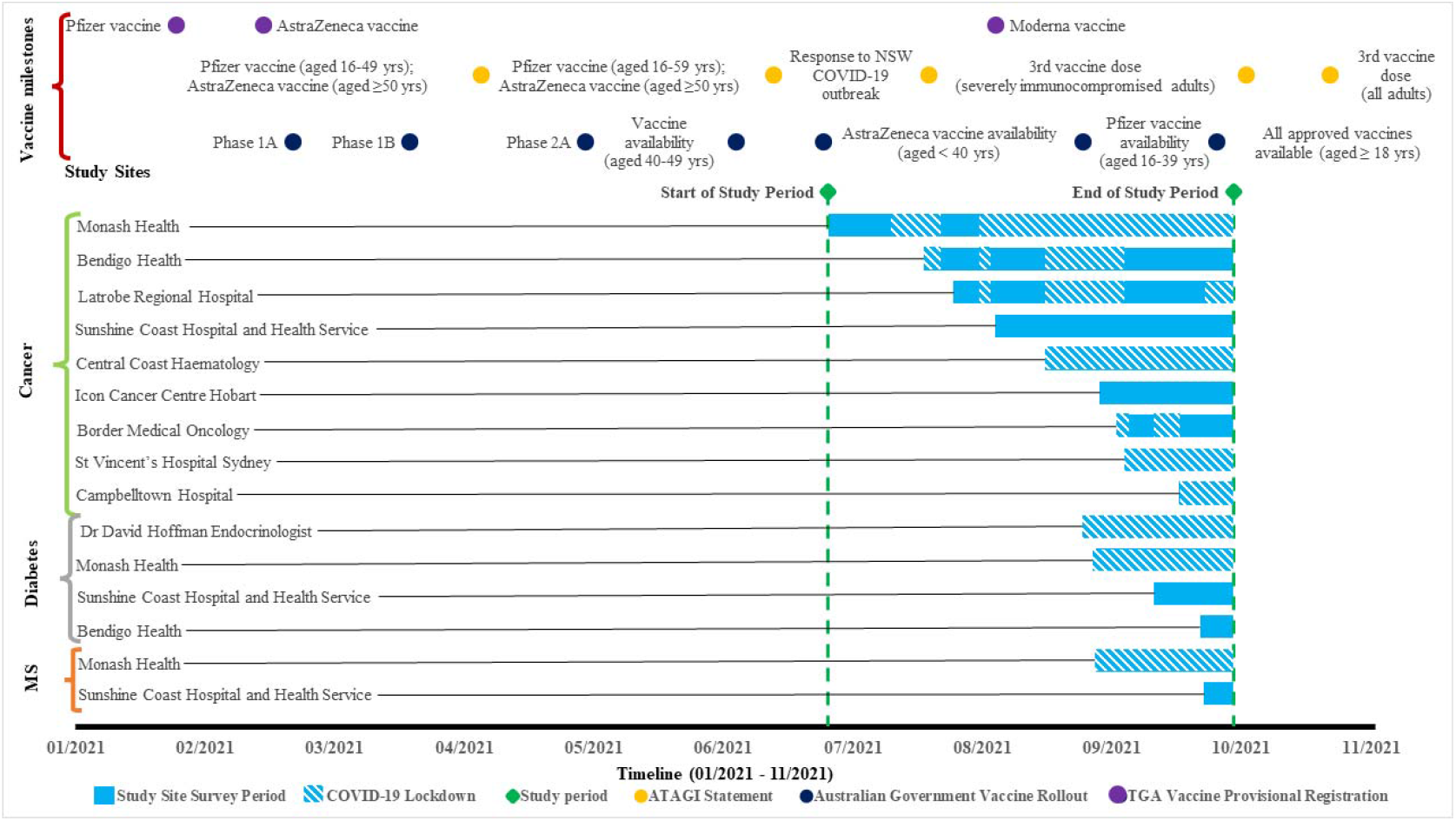
Survey timeline for each health service and participant group, with Australian State Government COVID-19 lockdowns embedded in the study site survey period. Yrs = years; MS = multiple sclerosis; ATAGI = Australian Technical Advisory Group on Immunisation; TGA = Therapeutic Goods Administration. Australian Government Vaccine Rollout Phase population group eligibility: Phase 1A rollout = Quarantine and border workers, health care workers, aged and disability residents and staff; Phase 1B rollout = Adults aged 70 years and over, Aboriginal and Torres Strait Islander people aged 55 years and over, Adults with underlying medical conditions, other critical and high-risk workers; Phase 2A rollout = Adults aged 50 years and over, Aboriginal and Torres Strait Islander people aged 18 years and over.

### Study participants

Participants were eligible if they had a past or current diagnosis of a solid organ or haematological malignancy, diabetes or MS; were aged 18 years and over; and a patient of a participating health service. People with cancer were recruited across nine healthcare services; people with diabetes were recruited at three sites and people with MS at two sites.

People with cancer and diabetes who had appointments scheduled in the next six months, and people with MS who attended an appointment in the previous 12 months, were invited to participate via text message invitation (one initial and a maximum of two follow-up messages) containing a link to participant information and electronic consent. Potential participants with cancer were also invited at their consultations and treatment appointments, and promotional materials were displayed at the health services.

Potential participants who accessed the survey link and gave informed consent were directed to the survey, hosted on the Qualtrics® secure data capture platform. The survey was presented in English.

### Measures

The survey was developed by a panel of clinicians, researchers and patient representatives (total 42 items for cancer and 44 items for diabetes and MS; Supplementary Table S1). All scale items used a 5-point Likert scale, plus a ‘don’t know’ option. No identifiable information was collected.

#### Vaccine uptake status, demographics, and clinical history

Demographic factors were collected including gender, age, level of highest education, range of household income, status as an Aboriginal and/or Torres Strait Islander person, and whether English was the participant’s first language. Clinical history items included disease type, time since diagnosis (within a range), and current treatment.

#### Oxford COVID-19 Vaccine Hesitancy Scale

This seven-item scale measures willingness to receive a COVID-19 vaccine, with higher scores indicating greater hesitancy. The scale was validated in a UK general population sample of 5114 adults, demonstrating excellent internal consistency (Cronbach’s alpha of 0.95).^9^ We adapted some items to maximise relevance to the Australian population and reflect the timing being post vaccine development. Specifically, reference to UK approval was removed; the first option was changed to ‘definitely/have taken’ in item 1; ‘when’ was substituted for ‘if’ in item 2; and ‘local area’ substituted for ‘local pharmacy’ in item 4 (supplementary information).

#### Oxford COVID-19 Vaccine Confidence and Complacency Scale

This 14-item scale measures confidence and complacency using four factors: collective importance of a vaccine, belief that COVID-19 infection may occur and the vaccine will work, speed of vaccine development, and side effects (Cronbach’s alpha range, 0.70 to 0.84).^9^ We adapted 11 of the 14 items by adding the words ‘I think’ to the beginning of the question. Higher scores indicated more negative attitudes toward vaccination (supplementary information).

#### Disease Influenced Vaccine Acceptance Scale-Six (DIVAS-6)

This scale was shown to evaluate aspects of vaccine-associated attitudes emanating from concerns about the patient’s disease and treatment (manuscript under review). The Disease Complacency indices, consisting of the first three items, were reverse scored so that higher scores indicate greater vaccine complacency. Higher scores in the Vaccine Vulnerability indices indicate greater perceived disease-related vaccine concerns.

### Statistical Analysis

Summary scores were calculated for each scale and subscale scores were calculated for each factor for the Oxford COVID-19 Vaccine Confidence and Complacency Scale and the DIVAS-6. ‘Don’t know’ responses were not scored or analysed, consistent with previous approaches.^9^ Data cleaning was performed to remove unsubmitted incomplete, duplicate and ineligible responses. Missing data were not imputed.

Socio-demographic, clinical data, and summary and subscale scores were summarised using descriptive statistics. Variables with too few classifications were combined or removed for analysis: this was required for no formal education level and primary education level (combined with secondary as highest education) and non-binary/other gender (removed for analysis due to statistical limitations because of the small number of observations).

Differences in demographics and individual scale items (item five ‘intent to vaccinate’ and item 12 ‘likelihood of COVID-19 infection’) between chronic disease type and vaccine uptake (any versus none) were detected with independent sample t-tests and chi-squared tests. Logistic regression was used to determine whether the scales (summary score, subscale score and items) predicted vaccination status. Linear regression was used to assess whether the Oxford scales’ summary and subscale scores predicted DIVAS-6 scores. Time since study commencement and demographic and disease-related variables that were significantly correlated with the outcome variable of interest and had a correlation of r >.10 (using Pearson’s and Spearman’s Rho) were controlled for in hierarchical regression analysis (Table 2). P-values <.05 were considered significant. Effect sizes were calculated with the phi coefficient (φ) and Cramér’s V (φ*c*) for chi-squared tests, and eta squared (η2) for independent sample t-tests. Statistical analyses were performed using SPSS Statistics Version 27.0 (IBM, USA). Findings from this study were reported according to STROBE (strengthening the reporting of observational studies in epidemiology) guidelines.^20^

### Patient and public involvement

The research team included a consumer advocate and consumer researcher who were both involved in design of the reported data by providing input on survey items and recruitment approaches and testing the survey, as well as interpretation of results and manuscript drafting and review.

## Results

### Participant characteristics

Of 8232 responses, unsubmitted incomplete (n = 3037), duplicate (n = 47), and ineligible (n = 465) responses were removed. There were 4683 eligible responses, from 3560 people with cancer, 842 people with diabetes and 281 people with MS. Socio-demographic characteristics are detailed in Table 1. Consistent with disease distribution, the proportion of females with MS was significantly higher than for cancer and diabetes, and there was a lower mean participant age. There were fewer university educated participants, a lower income distribution and greater proportion of participants who reported English as a second language in the diabetes cohort.

**Table 1.** Participant characteristics.

| Characteristic | All<br>N = 4683*<br>n, (%) | Cancer<br>n = 3560<br>n, (%) | Diabetes<br>n = 842<br>n, (%) | MS<br>n = 281<br>n, (%) | X<sup>2</sup> p-value | Phi coefficient |
| --- | --- | --- | --- | --- | --- | --- |
| **Gender** |  |  |  |  | <.001 | .14 |
| Male | 2108 (45.0) | 1586 (44.5) | 457 (54.3) | 65 (23.1) |  |  |
| Female | 2548 (54.4) | 1957 (55.0) | 378 (44.9) | 213 (75.8) |  |  |
| Non-Binary/Other | 27 (0.6) | 17 (0.5) | 7 (0.8) | 3 (1.1) |  |  |
| **Age** |  |  |  |  | <.001 | .03<sup>a</sup> |
| Mean (SD) | 60.6 (13.3) | 63.0 (12.0) | 55.1 (14.6) | 47.7 (12.8) |  |  |
| 18 – 29 | 119 (2.5) | 25 (0.7) | 65 (7.7) | 29 (10.3) |  |  |
| 30 - 49 | 793 (16.9) | 485 (13.6) | 186 (22.1) | 122 (43.4) |  |  |
| 50 - 69 | 2439 (52.1) | 1864 (52.4) | 455 (54.0) | 120 (42.7) |  |  |
| ≥70 | 1328 (28.4) | 1182 (33.2) | 136 (16.2) | 10 (3.6) |  |  |
| **Highest level of education** |  |  |  |  | <.001 | .06<sup>a</sup> |
| No formal / Primary | 131 (2.8) | 90 (2.5) | 37 (4.4) | 4 (1.4) |  |  |
| Secondary | 1572 (33.6) | 1167 (32.8) | 319 (37.9) | 86 (30.6) |  |  |
| Vocational/Trade | 1202 (25.6) | 896 (25.2) | 231 (27.4) | 75 (26.7) |  |  |
| University | 1764 (37.7) | 1396 (39.2) | 253 (30.0) | 115 (40.9) |  |  |
| Other | 14 (0.3) | 11 (0.3) | 2 (0.2) | 1 (0.4) |  |  |
| **Annual Household Income (AUD)** |  |  |  |  | <.001 | .07<sup>a</sup> |
| <50,000 | 1550 (33.1) | 1155 (32.4) | 323 (38.4) | 72 (25.6) |  |  |
| 50,000 – 100,000 | 1147 (24.5) | 854 (24.0) | 217 (25.8) | 76 (27.0) |  |  |
| 100,000 – 150,000 | 595 (12.7) | 462 (13.0) | 92 (10.9) | 41 (14.6) |  |  |
| >150,000 | 556 (11.9) | 464 (13.0) | 52 (6.2) | 40 (14.2) |  |  |
| Prefer not to say | 835 (17.8) | 625 (17.6) | 158 (18.8) | 52 (18.5) |  |  |
| **Aboriginal / Torres Strait Islander** |  |  |  |  |  | .. |
| Yes | 77 (1.6) | 51 (1.4) | 25 (3.0) | 1 (0.4) |  |  |
| No | 4560 (97.4) | 3475 (97.6) | 805 (95.6) | 280 (99.6) |  |  |
| Prefer not to say | 46 (1.0) | 34 (1.0) | 12 (1.4) | 0 (0.0) |  |  |
| **English as primary language** |  |  |  |  | <.001 | .13 |
| Yes | 4174 (89.1) | 3240 (91.0) | 677 (80.4) | 257 (91.5) |  |  |
| No | 508 (10.8) | 319 (9.0) | 165 (19.6) | 24 (8.5) |  |  |
| **Location** |  |  |  |  | <.001 | .10 |
| Metropolitan location | 3234 (69.1) | 2390 (67.1) | 605 (71.9) | 239 (85.1) |  |  |
| Regional/Rural location | 1449 (30.9) | 1170 (32.9) | 237 (28.1) | 42 (14.9) |  |  |
| **Vaccination Status** |  |  |  |  | .45 | .02 |
| Vaccinated | 3813 (81.5) | 2884 (81.1) | 696 (82.7) | 233 (82.9) |  |  |
| Unvaccinated | 868 (18.5) | 2884 (18.9) | 146 (17.3) | 48 (17.1) |  |  |
| *Exact cohort Age n = 4680; English as a first language n = 4682; vaccination status n=4681 (2 with missing data in the cancer group).<br><sup>a</sup>Cramer's V reported.<br>Chi-square for gender did not include non-binary/other, highest educational level did not include other, and was not undertaken for Aboriginal/Torres Strait Islander status, due to limited observations. X<sup>2</sup>= Chi-square; AUD = Australian Dollars. Vaccinated was defined as receiving at least one COVID-19 vaccine dose. |  |  |  |  |  |  |

Among participants with cancer, the most common types were breast (27.7%), haematological (24.4%), genitourinary (14.1%), gastrointestinal (13.8%) and lung (7.0%). Forty-four percent had been diagnosed with cancer in the last two years whereas 24.7% were diagnosed more than five years previously; 50.7% were receiving anti-cancer treatment at the time of survey completion.

Most participants with diabetes reported having type 2 (66.2%), while 29.9% reported type 1 and 3.9% reported other/don’t know. The majority (62.4%) had received their diagnosis more than ten years prior; 98.1% were receiving treatment including insulin (33.1%), non-insulin antidiabetic agents (21.9%) or a combination (43.2%).

Of participants with MS, 72.2% reported to have the relapsing-remitting type, 10.7% secondary-progressive; 9.3% primary-progressive and 7.8% other/don’t know. Just over half (52.0%) had received their diagnosis within the preceding ten years, and 79.7% were receiving current MS treatment.

### Vaccination status, intent and hesitancy

Overall, 81.5% of participants reported having at least one COVID-19 vaccine, comparable to vaccination prevalence in the total Australian population at the corresponding time point (79.9% had received at least one dose by October 4 2021).^21^ No significant difference in vaccine uptake between disease types was found. Unvaccinated status was significantly associated with decreased age, female sex, lower education and income levels, English as a second language, Aboriginal and Torres Strait Islander status, and residence in regional areas, although effect sizes were small (Table S2).

Ninety percent of participants reported they had or would definitely/probably accept a COVID-19 vaccine, 5.8% were unsure and 4.3% stated they were unlikely to accept a vaccine. The intent to be vaccinated did not vary significantly between disease types. As expected, there was a significant relationship between vaccine status and intent, with more vaccinated than unvaccinated participants reporting a likelihood to accept vaccination (definitely/ have taken or probably) (Table 2). Of participants who were unvaccinated, 52.0% stated that they were likely and 22.7% unlikely to accept a COVID-19 vaccine, while 25.3% were unsure. Participants who were unvaccinated had significantly higher total mean scores on the Oxford COVID-19 Hesitancy Scale -indicating greater vaccine hesitancy - compared to vaccinated participants, irrespective of disease type; this remained significant after controlling for covariates (Table 2).

**Table 2.**
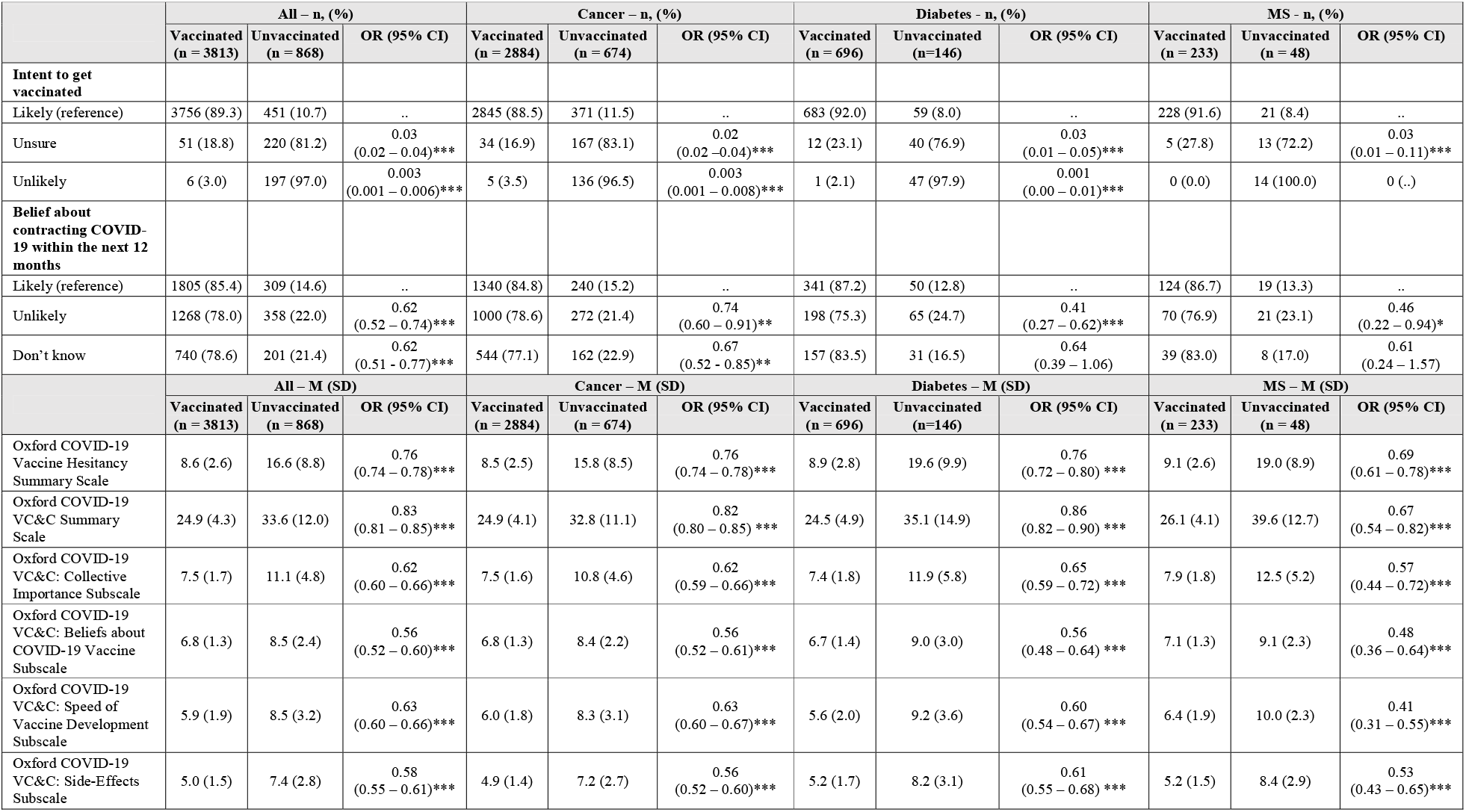
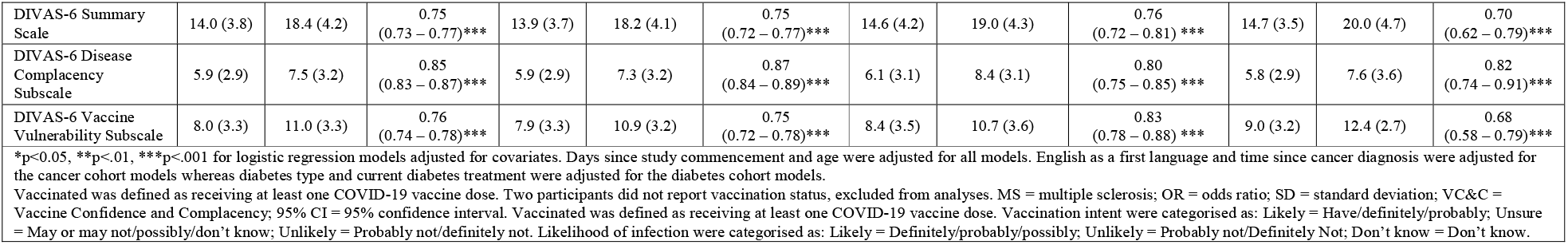
The relationship between vaccinated status with intent to get vaccinated, belief about contracting COVID-19 within the next 12 months, and the scale and subscale indices.

|  | All – n, (%) |  |  | Cancer – n, (%) |  |  | Diabetes – n, (%) |  |  | MS – n, (%) |  |  |
| --- | --- | --- | --- | --- | --- | --- | --- | --- | --- | --- | --- | --- |
|  | Vaccinated<br>(n = 3813) | Unvaccinated<br>(n = 868) | OR (95% CI) | Vaccinated<br>(n = 2884) | Unvaccinated<br>(n = 674) | OR (95% CI) | Vaccinated<br>(n = 696) | Unvaccinated<br>(n=146) | OR (95% CI) | Vaccinated<br>(n = 233) | Unvaccinated<br>(n = 48) | OR (95% CI) |
| **Intent to get vaccinated** |  |  |  |  |  |  |  |  |  |  |  |  |
| Likely (reference) | 3756 (89.3) | 451 (10.7) | .. | 2845 (88.5) | 371 (11.5) | .. | 683 (92.0) | 59 (8.0) | .. | 228 (91.6) | 21 (8.4) | .. |
| Unsure | 51 (18.8) | 220 (81.2) | 0.03<br>(0.02 – 0.04)*** | 34 (16.9) | 167 (83.1) | 0.02<br>(0.02 – 0.04)*** | 12 (23.1) | 40 (76.9) | 0.03<br>(0.01 – 0.05)*** | 5 (27.8) | 13 (72.2) | 0.03<br>(0.01 – 0.11)*** |
| Unlikely | 6 (3.0) | 197 (97.0) | 0.003<br>(0.001 – 0.006)*** | 5 (3.5) | 136 (96.5) | 0.003<br>(0.001 – 0.008)*** | 1 (2.1) | 47 (97.9) | 0.001<br>(0.00 – 0.01)*** | 0 (0.0) | 14 (100.0) | 0 (..) |
| **Belief about contracting COVID-19 within the next 12 months** |  |  |  |  |  |  |  |  |  |  |  |  |
| Likely (reference) | 1805 (85.4) | 309 (14.6) | .. | 1340 (84.8) | 240 (15.2) | .. | 341 (87.2) | 50 (12.8) | .. | 124 (86.7) | 19 (13.3) | .. |
| Unlikely | 1268 (78.0) | 358 (22.0) | 0.62<br>(0.52 – 0.74)*** | 1000 (78.6) | 272 (21.4) | 0.74<br>(0.60 – 0.91)** | 198 (75.3) | 65 (24.7) | 0.41<br>(0.27 – 0.62)*** | 70 (76.9) | 21 (23.1) | 0.46<br>(0.22 – 0.94)* |
| Don't know | 740 (78.6) | 201 (21.4) | 0.62<br>(0.51 – 0.77)*** | 544 (77.1) | 162 (22.9) | 0.67<br>(0.52 – 0.85)** | 157 (83.5) | 31 (16.5) | 0.64<br>(0.39 – 1.06) | 39 (83.0) | 8 (17.0) | 0.61<br>(0.24 – 1.57) |
|  | All – M (SD) |  |  | Cancer – M (SD) |  |  | Diabetes – M (SD) |  |  | MS – M (SD) |  |  |
|  | Vaccinated<br>(n = 3813) | Unvaccinated<br>(n = 868) | OR (95% CI) | Vaccinated<br>(n = 2884) | Unvaccinated<br>(n = 674) | OR (95% CI) | Vaccinated<br>(n = 696) | Unvaccinated<br>(n=146) | OR (95% CI) | Vaccinated<br>(n = 233) | Unvaccinated<br>(n = 48) | OR (95% CI) |
| Oxford COVID-19 Vaccine Hesitancy Summary Scale | 8.6 (2.6) | 16.6 (8.8) | 0.76<br>(0.74 – 0.78)*** | 8.5 (2.5) | 15.8 (8.5) | 0.76<br>(0.74 – 0.78)*** | 8.9 (2.8) | 19.6 (9.9) | 0.76<br>(0.72 – 0.80) *** | 9.1 (2.6) | 19.0 (8.9) | 0.69<br>(0.61 – 0.78)*** |
| Oxford COVID-19 VC&C Summary Scale | 24.9 (4.3) | 33.6 (12.0) | 0.83<br>(0.81 – 0.85)*** | 24.9 (4.1) | 32.8 (11.1) | 0.82<br>(0.80 – 0.85) *** | 24.5 (4.9) | 35.1 (14.9) | 0.86<br>(0.82 – 0.90) *** | 26.1 (4.1) | 39.6 (12.7) | 0.67<br>(0.54 – 0.82)*** |
| Oxford COVID-19 VC&C: Collective Importance Subscale | 7.5 (1.7) | 11.1 (4.8) | 0.62<br>(0.60 – 0.66)*** | 7.5 (1.6) | 10.8 (4.6) | 0.62<br>(0.59 – 0.66)*** | 7.4 (1.8) | 11.9 (5.8) | 0.65<br>(0.59 – 0.72) *** | 7.9 (1.8) | 12.5 (5.2) | 0.57<br>(0.44 – 0.72)*** |
| Oxford COVID-19 VC&C: Beliefs about COVID-19 Vaccine Subscale | 6.8 (1.3) | 8.5 (2.4) | 0.56<br>(0.52 – 0.60)*** | 6.8 (1.3) | 8.4 (2.2) | 0.56<br>(0.52 – 0.61)*** | 6.7 (1.4) | 9.0 (3.0) | 0.56<br>(0.48 – 0.64) *** | 7.1 (1.3) | 9.1 (2.3) | 0.48<br>(0.36 – 0.64)*** |
| Oxford COVID-19 VC&C: Speed of Vaccine Development Subscale | 5.9 (1.9) | 8.5 (3.2) | 0.63<br>(0.60 – 0.66)*** | 6.0 (1.8) | 8.3 (3.1) | 0.63<br>(0.60 – 0.67)*** | 5.6 (2.0) | 9.2 (3.6) | 0.60<br>(0.54 – 0.67) *** | 6.4 (1.9) | 10.0 (2.3) | 0.41<br>(0.31 – 0.55)*** |
| Oxford COVID-19 VC&C: Side-Effects Subscale | 5.0 (1.5) | 7.4 (2.8) | 0.58<br>(0.55 – 0.61)*** | 4.9 (1.4) | 7.2 (2.7) | 0.56<br>(0.52 – 0.60)*** | 5.2 (1.7) | 8.2 (3.1) | 0.61<br>(0.55 – 0.68) *** | 5.2 (1.5) | 8.4 (2.9) | 0.53<br>(0.43 – 0.65)*** |
| DIVAS-6 Summary Scale | 14.0 (3.8) | 18.4 (4.2) | 0.75<br>(0.73 – 0.77)*** | 13.9 (3.7) | 18.2 (4.1) | 0.75<br>(0.72 – 0.77)*** | 14.6 (4.2) | 19.0 (4.3) | 0.76<br>(0.72 – 0.81) *** | 14.7 (3.5) | 20.0 (4.7) | 0.70<br>(0.62 – 0.79)*** |
| DIVAS-6 Disease Complacency Subscale | 5.9 (2.9) | 7.5 (3.2) | 0.85<br>(0.83 – 0.87)*** | 5.9 (2.9) | 7.3 (3.2) | 0.87<br>(0.84 – 0.89)*** | 6.1 (3.1) | 8.4 (3.1) | 0.80<br>(0.75 – 0.85) *** | 5.8 (2.9) | 7.6 (3.6) | 0.82<br>(0.74 – 0.91)*** |
| DIVAS-6 Vaccine Vulnerability Subscale | 8.0 (3.3) | 11.0 (3.3) | 0.76<br>(0.74 – 0.78)*** | 7.9 (3.3) | 10.9 (3.2) | 0.75<br>(0.72 – 0.78)*** | 8.4 (3.5) | 10.7 (3.6) | 0.83<br>(0.78 – 0.88) *** | 9.0 (3.2) | 12.4 (2.7) | 0.68<br>(0.58 – 0.79)*** |
| *p<0.05, **p<.01, ***p<.001 for logistic regression models adjusted for covariates. Days since study commencement and age were adjusted for all models. English as a first language and time since cancer diagnosis were adjusted for the cancer cohort models whereas diabetes type and current diabetes treatment were adjusted for the diabetes cohort models.<br>Vaccinated was defined as receiving at least one COVID-19 vaccine dose. Two participants did not report vaccination status, excluded from analyses. MS = multiple sclerosis; OR = odds ratio; SD = standard deviation; VC&C = Vaccine Confidence and Complacency; 95% CI = 95% confidence interval. Vaccinated was defined as receiving at least one COVID-19 vaccine dose. Vaccination intent were categorised as: Likely = Have/definitely/probably; Unsure = May or may not/possibly/don't know; Unlikely = Probably not/definitely not. Likelihood of infection were categorised as: Likely = Definitely/probably/possibly; Unlikely = Probably not/Definitely Not; Don't know = Don't know. |  |  |  |  |  |  |  |  |  |  |  |  |

### Vaccine-related beliefs (Oxford COVID-19 Confidence and Complacency Scale)

Participants who were unvaccinated scored higher on the Oxford COVID-19 Confidence and Complacency Scale and subscales (Table 2), signifying unvaccinated status was associated with more negative attitudes toward COVID-19 vaccines, including beliefs regarding collective importance, potential therapeutic benefits, speed of development and side effects. Unvaccinated respondents were significantly less likely to believe they would become infected with SARS-CoV-2 in the next 12 months compared to those who were vaccinated (B = - 0.48, p<.001, OR CI 0.52 – 0.74). More negative attitudes toward COVID-19 vaccines were strongly correlated with greater vaccine hesitancy (Oxford COVID-19 Hesitancy Scale), Pearson’s r = 0.78, n = 2748, p<0.001.

### Impact of underlying disease (DIVAS-6)

Of all participants, 60.6% reported they were worried (strongly agree or somewhat agree) about COVID-19 infection and 69.9% felt that the vaccine was more important to them due to their underlying disease. Eighty percent endorsed that physician recommendations regarding the vaccine were important to them. Forty-four percent of respondents were concerned about vaccine efficacy due to their underlying disease, while 39.6% and 25.7% respectively reported concerns of vaccine effect on their disease or treatment (Figure S1).

Positive relationships were observed between the DIVAS-6 total and subscale scores and the two Oxford scales of COVID-19 Vaccine Hesitancy and Vaccine Confidence and Complacency, demonstrating convergent validity of the DIVAS-6 with other measures of vaccine hesitancy and attitudes. Overall, unvaccinated participants had higher DIVAS-6 total and subscale scores, indicating greater complacency about potential COVID-19 infection and greater concerns of vaccine efficacy and the impact of vaccination on their disease and/or treatment (Table 2).

## Discussion

### Principal findings

In a large cohort of patients with serious and/or chronic morbidity, we found that underlying disease significantly affected attitudes and uptake of COVID-19 vaccination. Interestingly, the level and type of concerns were common across the three major diseases studied. While most participants acknowledged increased apprehension about SARS-CoV-2 infection and the importance of the vaccine due to underlying disease, between a quarter-to-half were worried about vaccine efficacy and/or the effect of the vaccine on their disease. To our knowledge, this is the first study in a medically vulnerable population to use this suite of validated instruments, including a scale to specifically assess disease-related influences on vaccine acceptance. Alongside disease-related concerns, we observed similar explanatory factors of vaccine acceptance to that seen in the general public,^4 5 9 22^ including collective importance and sociodemographic characteristics.

### Findings in context

Our study measured concurrently vaccine uptake and hesitancy. The prevalence of vaccine uptake in our cohort was slightly higher than the general population at the time of the survey.^21^ Not surprisingly, given their intrinsic vulnerability, we found low vaccine hesitancy, compared to the rate in the general population at the time.^10^ The level was consistent with other studies in populations with comorbidities,^18 22 23^ although several have reported higher hesitancy levels.^6 24^ This may be explained by differences in survey timing, geographic region and the definition and scales of hesitancy used.

Vaccine hesitancy is viewed as a continuum which may change with time and co-exist with vaccine compliance. The phenomenon of the ‘vaccinated but hesitant’ is well described.^25^ In one US study, 60% of recently COVID-19 vaccinated individuals expressed some level of hesitancy.^26^ Conversely, motivations toward vaccination may not convert to vaccine uptake. One in five participants in our study remained unvaccinated, even though over half of this cohort expressed a positive intent to pursue vaccination. Incongruity between attitude and behaviour point to potential underlying concerns, leading to ambivalence or delay. Practical considerations, such as barriers to access, may also be responsible although in Australia this was largely mitigated by this ‘co-morbidity’ group having prioritised free access to vaccines (available for around six months by end of survey period).

Clinical decision-making in people with chronic disease is acknowledged to be a multi-layered process, taking into consideration clinical factors, personal values and sociocultural influences.^27^ Similarly, drivers of any vaccine hesitancy are likely to be complex and context dependent. Our study clearly shows that disease-related considerations were prominent in medically vulnerable populations and had influenced COVID-19 vaccination decision-making. Despite confirmation of vaccine safety in multiple studies in high risk populations without detrimental effect on underlying disease,^15 22^ concerns about vaccine effect on disease stability and interactions with current treatment were commonly cited as reasons for vaccine non-acceptance in our study, supporting the findings of smaller studies in cancer and autoimmune disease.^6 18 19 28^ The high prevalence of vaccine-related efficacy and safety concerns, even in a highly vaccinated and vaccine-accepting population is worthy of noting, particularly with growing data supporting the requirement for booster doses.^15 16^ This underscores a significant information gap for people with underlying illnesses, especially addressing the interaction of COVID-19 vaccination with their disease course and therapy.

### Implications for public health policy and practice

To engage medically vulnerable populations, targeted interventions by trusted sources are likely to be crucial and will complement public health campaigns which have largely focussed on broad societal benefits of vaccination. Clinically relevant, evidence-based data need to be delivered promptly to medically vulnerable populations using consistent and simple messaging to alleviate concerns and any misconceptions, consolidate positive attitudes and build confidence in vaccination strategies and health care services. Although there have been information campaigns, often produced by Non-Government Organisations (e.g. the U.S. National Multiple Sclerosis Society provides a succinct unambiguous expert-reviewed COVID-19 guidance to address the rationale for vaccination, as well as safety aspects in the context of MS and disease modifying therapy),^29^ there is a need for centralisation to ensure consistency and maintain updates as new information emerges. The risk of outdated information, in a pandemic with rapidly evolving variants that have different degrees of protection from current vaccines, needs to be mitigated.

Educational programs for health care providers are also imperative, based on the high proportion of our participants who valued their physician’s recommendations on COVID-19 vaccines, in line with other studies.^6 18 28^ To date however, there is a paucity of formal guidance and specific training for clinicians, manifesting as patients reporting receiving conflicting advice from different clinicians.^30^ The DIVAS-6 scale was designed to be easily administered at the point of care to facilitate discussions directed to individual concerns. People who score highly on its Disease Complacency subscale may benefit from specific education on the increased risk of COVID-19 for people with serious illnesses, while those who score highly on the Vaccine Vulnerability subscale may be assisted by education on the safety aspects of the vaccine.

### Strengths and limitations

In addition to the use of validated instruments, the strengths of the present study include the broad and diverse sampling of pre-defined, high-risk participants with three disease types within a range of demographics. The three-month recruitment period spanned changing levels of community transmission and social restriction measures, including strict state-wide lockdowns, giving a broader perspective than a short ‘snapshot’ survey. Importantly, data collection commenced at least three months after the availability of vaccines to high-risk groups, allowing sufficient time for vaccination if willing. Finally, the low prevalence of COVID-19 disease in Australia at the time of the study gives insight into attitudes and actions in the setting where there is less immediate threat from COVID-19.

Study limitations include recall and misclassification bias inherent in a survey-based study and availability only in English. Additionally, people who had a greater interest in COVID-19 vaccinations may have been more likely to respond. These issues should be considered when generalising findings. Given that Australia had one of the highest vaccination rates globally and relatively low community transmission during most of the period studied, results may not be directly translatable to global regions with lower vaccination rates and/or high transmission. However, within one year of the study, a period of extremely high community transmission occurred throughout Australia and with now well-established patterns in this pandemic, understanding attitudes at times of lower community transmission is probably as important as at times of high prevalence. Determinants of vaccine behaviour are likely to evolve over time in the context of changing disease epidemiology, therapeutic options and vaccine characteristics; serial studies are required to assess these dynamics.

### Conclusions

In a large Australian cross-sectional study in people with cancer, diabetes and MS, we found high rates of COVID-19 vaccination uptake and positive intent, that importantly, seemed to be influenced by concerns of the impact of SARS-CoV-2 infection on their underlying disease. These concerns were both for the need for vaccination to protect them due to their inherent vulnerability arising from their disease and/or associated treatment, as well as for the potential impact of vaccination on their ability to maintain control of their illness. Our findings highlight the importance of addressing the concerns of medically vulnerable populations, including actively targeted public health communication strategies and health professional training, to optimise COVID-19 vaccine acceptance.

## Data Availability

All data produced in the present study are available upon reasonable request to the authors.

