## Supplementary materials for "Serious underlying medical conditions and COVID-19 vaccine hesitancy"

This appendix has been provided by the authors to give readers additional information about their work

### List of investigators

| **Study Site, Australian State** | **Investigator(s)** |
| --- | --- |
| **Monash Health, Victoria** |  |
| CANVACCS | Eva Segelov |
|  | Daphne Day |
|  | Lisa Grech |
|  | Mike Nguyen |
|  | Nathan Bain |
|  | Amelia McCartney |
|  | Kate Webber |
|  | Elizabeth Ahern |
|  | Muhammad Alamgeer |
|  | Amy Body |
|  | Peter Briggs |
|  | Sophia Frentzas |
|  | Marion Harris |
|  | Gwo-Yaw Ho |
|  | Caroline Lum |
|  | Cameron McLaren |
|  | David Pook |
|  | Andrew Strickland |
|  | Michelle White |
|  | Vi Luong |
|  | Avraham Travers |
|  | Walid Zwieky |
|  | Veronica Lopez Aedo |
| DIABVACCS | Jennifer Wong |
|  | Barbora de Courten |
| MSVACCS | Ernest Butler |
|  | Michelle Allan |
| **Bendigo Health, Victoria** |  |
| CANVACCS | Sam Harris |
| DIABVACCS | Mark Savage |
|  | Frank Gao |
|  | Amy Harding |
| **Latrobe Regional Hospital, Victoria** |  |
| CANVACCS | Hieu Chau |
| **Sunshine Coast Hospital and Health Service, Queensland** |  |
| CANVACCS | Bryan Chan |
| DIABVACCS | Brett Sillars |
| MSVACCS | Antony Winkel |
|  | Joshua Barton |
| **Icon Cancer Centre Hobart, Tasmania** |  |
| CANVACCS | Louise Nott |
| **Central Coast Haematology, New South Wales** |  |
| CANVACCS | Richard Blennerhassett |
|  | Cecily Forsyth |
|  | Jacqueline Jagger |
| **St Vincent’s Hospital Sydney, New South Wales** |  |
| CANVACCS | Nada Hamad |
| **Campbelltown Hospital, New South Wales** |  |
| CANVACCS | Annette Tognela |
| **Dr David Hoffman, New South Wales** |  |
| DIABVACCS | David Hoffman |
| **Border Medical Oncology, New South Wales** |  |
| CANVACCS | Craig Underhill |
| CANVACCS = CANcer patients’ perspectives on coronavirus VACCination Survey  DIABVACCS = DIABetes patients’ perspectives on coronavirus VACCination Survey  MSVACCS = Multiple Sclerosis patients’ perspectives on coronavirus VACCination Survey | |

### Table S1. Chronic disease COVID-19 Vaccine Survey Items.

| **Screening items** | |
| --- | --- |
| 1. Are you 18 years or older? |  Yes   No (Terminate if No) |
| 2. Have you got [chronic disease]? |  Yes   No (Terminate if No) |
| 3. Are you a [participating site] patient? |  Yes   No (Terminate if No) |
| **Vaccination status** | |
| 4. Have you already received a COVID-19 vaccine? |  Yes, 1 dose only   Yes, 2 doses   No |
| **Oxford COVID-19 Vaccine Hesitancy Scale**  Instructions: We would like to know your feelings and thoughts about the COVID-19 vaccine. Note: If you have already been vaccinated against COVID-19, please complete these questions in relation to a future COVID-19 vaccine dose/booster. | |
| 5. Would you take a COVID-19 vaccine if offered? |  Definitely/have taken   Probably   I may or may not   Probably not   Definitely not   Don't know |
| 6. When a COVID-19 vaccine is available: |  I will want to get it as soon as possible   I will take it when offered   I'm not sure what I will do   I will put off (delay) getting it   I will refuse to get it   Don't know |
| 7. I would describe my attitude towards receiving a COVID-19 vaccine as: |  Very keen   Pretty positive   Neutral   Quite uneasy   Against it   Don't know |
| 8. If a COVID-19 vaccine was available in my local area, I would: |  Get it as soon as possible   Get it when I have time   Delay getting it   Avoid getting it for as long as possible   Never get it   Don't know |
| 9. If my family or friends were thinking of getting a COVID-19 vaccination, I would: |  Strongly encourage them   Encourage them   Not say anything to them about it   Ask them to delay getting the vaccination   Suggest that they do not get the vaccination   Don't know |
| 10. I would describe myself as: |  Eager to get a COVID-19 vaccine   Willing to get the COVID-19 vaccine   Not bothered about getting the COVID-19 vaccine   Unwilling to get the COVID-19 vaccine   Anti-vaccination for COVID-19   Don't know |
| 11. Taking a COVID-19 vaccination is: |  Really important   Important   Neither important nor unimportant   Unimportant   Really unimportant   Don’t know |
| **Oxford COVID-19 Vaccine Confidence and Complacency Scale** | |
| 12. Do you think you will be infected with COVID-19 over the next 12 months? |  Definitely   Probably   Possibly   Probably not   Definitely not   Don’t know |
| 13. I think the COVID-19 vaccine is likely to: |  Work for almost everyone   Work for most people   I am unsure how many people it will work for   Not work for most people   Not work for anyone   Don’t know |
| 14. I think the COVID-19 vaccine is likely to: |  Definitely work for me   Probably work for me   May or may not work for me   Probably not work for me   Definitely not work for me   Don’t know |
| 15. I think if I get the COVID-19 vaccine it will be: |  Really helpful for the community around me   Helpful for the community around me   Neither helpful nor unhelpful for the community around me   Unhelpful for the community around me   Really unhelpful for the community around me   Don’t know |
| 16. I think if individuals like me get the COVID-19 vaccine it will: |  Save a large number of lives   Save some lives   Have no impact   Lead to more deaths   Lead to a large number of deaths   Don’t know |
| 17. I think the speed of developing and testing the vaccine means it will be: |  Really good   Good   Will not affect how good or bad it is   Bad   Really bad   Don’t know |
| 18. I think the speed of developing and testing the vaccine means it will be: |  Really safe   Safe   It will not affect how safe it is   Unsafe   Really unsafe   Don’t know |
| 19. I think if many people do not get the vaccine this: |  Will be dangerous   May be dangerous   Will have no consequences at all   May be good   Will be good   Don’t know |
| 20. I expect that receiving the vaccine will be: |  Hardly noticeable   A little unpleasant   Moderately unpleasant   Painful   Extremely painful   Don’t know |
| 21. I think the side-effects for people of getting the COVID-19 vaccine will be: |  None   Mild   Moderate   Significant   Life-threatening   Don’t know |
| 22. I think the COVID-19 vaccine will: |  Greatly strengthen my immune system   Strengthen my immune system   It will neither strengthen nor weaken my immune system   Weaken my immune system   Greatly weaken my immune system   Don’t know |
| 23. I think taking the COVID-19 vaccine: |  Will give me complete freedom to get on with life just as before   Will give me greater freedom   Will have no effect on my freedom   Will restrict my freedom   Will completely restrict my freedom to get on with life   Don’t know |
| 24. I think getting the vaccine is a sign of: |  Great personal strength   Personal strength   Not a sign of personal strength or weakness   Personal weakness   Great personal weakness   Don’t know |
| 25. Taking a new COVID-19 vaccine will make me feel like a guinea pig: |  Do not agree   Agree a little   Agree moderately   Agree a lot   Completely agree   Don’t know |
| **Disease Influenced Visual Acceptance Scale-Six**  Instructions: We would like to know about how your [chronic disease] may be related to your feelings and thoughts about the COVID-19 vaccine. For each of the following statements, please tap/click the one choice that best represents how strongly you agree or disagree with it. There are 6 choices to choose from for each statement. | |
| 26. My history of [chronic disease] makes me more worried about being infected with COVID-19: |  Strongly agree   Somewhat agree   Neither disagree nor agree   Somewhat disagree   Strongly disagree   Don’t know |
| 27. My history of [chronic disease] means having the vaccine is more important to me: |  Strongly agree   Somewhat agree   Neither disagree nor agree   Somewhat disagree   Strongly disagree   Don’t know |
| 28. My doctor’s recommendation regarding the vaccine is important to me: |  Strongly agree   Somewhat agree   Neither disagree nor agree   Somewhat disagree   Strongly disagree   Don’t know |
| 29. My history of [chronic disease] makes me worried about how well the vaccine will work for me: |  Strongly disagree   Somewhat disagree   Neither disagree nor agree   Somewhat agree   Strongly agree   Don’t know |
| 30. My history of [chronic disease] makes me worried about how the vaccine will affect me: |  Strongly disagree   Somewhat disagree   Neither disagree nor agree   Somewhat agree   Strongly agree   Don’t know |
| 31. I am worried about how the vaccine will affect my [chronic disease] treatment: |  Strongly disagree   Somewhat disagree   Neither disagree nor agree   Somewhat agree   Strongly agree   Don’t know |
| **Demographics** | |
| 32. What is your gender? |  Male   Female   Non-binary / third gender   Prefer not to say |
| 33. What is your age? | _________________ |
| 34. What is your highest educational level (completed)? |  No formal education   Primary education   Secondary education   Vocational/trade qualification   University education or higher degree   Other (please specify): _____________________ |
| 35. What is your annual household income (including everyone who lives in your home)? |  Less than $50,000   $50,001 to $100,000   $100,001 to $150,000   More than $150,000   Prefer not to say |
| 36. Do you identify as Aboriginal and/or Torres Strait Islander? |  Yes   No   Prefer not to say |
| 37. Is English your first language? |  Yes   No |
| 38. Please include any comments about your feelings and thoughts about your [chronic disease] and COVID-19 vaccination that you would like to share. If you have no comments to include, please type 'Nil'. | [Text entry]: _______________________________________ |
| **Disease-specific clinical questions (Cancer only)** | |
| 39. What type of cancer do you have? (i.e. where your cancer started and not where it has spread to). |  Breast   Lung (including mesothelioma)   Genitourinary (i.e. prostate, kidney, testicular or bladder)   Skin (including melanoma)   Gastrointestinal (i.e. stomach, oesophagus, bile duct, gallbladder, pancreas, colon, rectum, or anus)   Gynaecological (i.e. ovarian, cervical, uterine or vulvar/vaginal)   Head and neck (i.e. mouth, throat, sinus or nose)   Brain   Blood (i.e. leukemia, myeloma and lymphoma)   Other (please specify):___________________________ |
| 40. When was your cancer diagnosed? |  Less than 6 months ago   6 to 24 months ago   2 to 5 years ago   More than 5 years ago |
| 41. As you understand it, is your cancer in just the one area where it started (localised) or has it spread to other places in the body (metastatic)? |  Localised   Metastatic   Don't know   Other (please type any comments): _____________________ |
| 42. Are you currently on cancer treatment? |  Yes   No |
| 43. How long ago was your last treatment? (e.g. chemotherapy, immunotherapy, hormonal treatment, targeted therapy, radiotherapy and/or clinical trial). |  Currently on treatment   Less than 1 year ago   1 to 5 years ago   More than 5 years ago |
| **Disease-specific clinical questions (Diabetes only)** | |
| 39. What type of diabetes do you have? |  Type 1   Type 2   Other (please specify): _________________________________   Don’t know |
| 40. How long have you had diabetes? |  Less than 1 year   1 to 5 years   5.1 to 10 years   More than 10 years |
| 41. My most recent HbA1c (within the past year) was: |  Less than 7%   7% to 8.5%   8.6% to 10%   More than 10%   Don't know |
| 42. My current treatment for my diabetes is/are (select all that apply): |  Insulin   Tablets   Diet only   Injectables (not insulin)   Other:  _________________________________ |
| 43. In the past month, would you say that your management of diabetes was: |  Excellent   Very good   Good   Fair   Poor |
| 44. In the last four weeks, how much did your diabetes affect your daily activities? |  All the time   Most of the time   Some of the time   Not very often   Not at all |
| **Disease-specific clinical questions (Multiple Sclerosis only)** | |
| 39. What type of MS do you have? |  Relapsing-remitting MS (RRMS)   Primary progressive MS (PPMS)   Secondary progressive MS (SPMS)   Other (please specify): ________________________________   Don't know |
| 40. How long have you had MS? |  Less than 1 year   1 to 5 years   5.1 to 10 years   More than 10 years |
| 41. My current treatment for my MS is/are (select all that apply): |  Tablets   Injectables   Intravenous   No specific treatment   Other: _______________________________________ |
| 42. Over the past 6 months, is your MS well controlled? |  Yes   No   Don’t know |
| 43. People often have difficulty taking their medications for one reason or another. How many times have you missed taking your disease modifying therapies (DMT) in the past month? |  All of the time   Most of the time   Some of the time   Occasionally   Never |
| 44. In the last four weeks, how much did your MS affect your daily activities? |  All the time   Most of the time   Some of the time   Not very often   Not at all |

### Table S2. Participant characteristics, by vaccination status and disease.

|  | **All diseases**  **n, (%)** | | | | **Cancer**  **n, (%)** | | | | **Diabetes**  **n, (%)** | | | | **MS**  **n, (%)** | | | |
| --- | --- | --- | --- | --- | --- | --- | --- | --- | --- | --- | --- | --- | --- | --- | --- | --- |
| **Characteristics** | **Vaccinated**  **(n = 3813)** | **Unvaccinated (n = 868)** | **p^a^** | **ϕ**^b^ | **Vaccinated**  **(n = 2884)** | **Unvaccinated (n = 674)** | **p^a^** | **ϕ**^b^ | **Vaccinated (n = 696)** | **Unvaccinated (n=146)** | **p^a^** | **ϕ**^b^ | **Vaccinated**  **(n = 233)** | **Unvaccinated**  **(n = 48)** | **p^a^** | **ϕ**^b^ |
| **Gender** |  |  | 0.002 | 0.05 |  |  | 0.004 | 0.05 |  |  | 0.37 | 0.03 |  |  | 0.35 | 0.07 |
| Male | 1758 (83.5) | 348 (16.5) |  |  | 1318 (83.2) | 266 (16.8) |  |  | 383 (83.8) | 74 (16.2) |  |  | 57 (87.7) | 8 (12.3) |  |  |
| Female | 2034 (79.8) | 514 (20.2) |  |  | 1553 (79.4) | 404 (20.6) |  |  | 307 (81.2) | 71 (18.8) |  |  | 174 (81.7) | 39 (18.3) |  |  |
| Non-Binary/Other | 21 (77.8) | 6 (22.2) |  |  | 13 (76.5) | 4 (23.5) |  |  | 6 (85.7) | 1 (14.3) |  |  | 2 (66.7) | 1 (33.3) |  |  |
| **Age** |  |  | <0.001^c^ | 0.03^c^ |  |  | <0.001^c^ | 0.04 ^c^ |  |  | <0.001^c^ | 0.03 ^c^ |  |  | <0.001^c^ | 0.06^c^ |
| Mean (SD) | 61.8 (13.0) | 55.6 (13.5) |  |  | 64.1 (11.7) | 57.9 (12.3) |  |  | 56.3 (14.3) | 49.8 (14.6) |  |  | 49.1 (12.6) | 40.6 (11.7) |  |  |
| 18 – 29 | 79 (66.4) | 40 (33.6) |  |  | 13 (52.0) | 12 (48.0) |  |  | 47 (72.3) | 18 (27.7) |  |  | 19 (65.5) | 10 (34.5) |  |  |
| 30 - 49 | 576 (72.6) | 217 (27.4) |  |  | 341 (70.3) | 144 (29.7) |  |  | 141 (75.8) | 45 (24.2) |  |  | 94 (77.0) | 28 (23.0) |  |  |
| 50 - 69 | 1953 (80.1) | 486 (19.9) |  |  | 1458 (78.2) | 406 (21.8) |  |  | 384 (84.4) | 71 (15.6) |  |  | 111 (92.5) | 9 (7.5) |  |  |
| ≥70 | 1202 (90.6) | 125 (9.4) |  |  | 1069 (90.5) | 112 (9.5) |  |  | 124 (91.2) | 12 (8.8) |  |  | 9 (90.0) | 1 (10.0) |  |  |
| **Highest level of education** |  |  | <0.001 | 0.07^d^ |  |  | <0.001 | 0.07^d^ |  |  | 0.37 | 0.06^d^ |  |  | 0.23 | 0.12^d^ |
| No formal/Primary | 96 (73.3) | 35 (26.7) |  |  | 66 (73.3) | 24 (26.7) |  |  | 27 (73.0) | 10 (27.0) |  |  | 3 (75.0) | 1 (25.0) |  |  |
| Secondary | 1272 (81.0) | 298 (19.0) |  |  | 931 (79.9) | 234 (20.1) |  |  | 268 (84.0) | 51 (16.0) |  |  | 73 (84.9) | 13 (15.1) |  |  |
| Vocational/Trade | 951 (79.1) | 251 (20.9) |  |  | 705 (78.7) | 191 (21.3) |  |  | 189 (81.8) | 42 (18.2) |  |  | 57 (76.0) | 18 (24.0) |  |  |
| University | 1487 (84.3) | 277 (15.7) |  |  | 1175 (84.2) | 221 (15.8) |  |  | 212 (83.8) | 41 (16.2) |  |  | 100 (87.0) | 15 (13.0) |  |  |
| Other | 7 (50.0) | 7 (50.0) |  |  | 7 (63.6) | 4 (36.4) |  |  | 0 (0.0) | 2 (100.0) |  |  | 0 (0.0) | 1 (100.0) |  |  |
| **Annual Household Income (AUD)** |  |  | <0.001 | 0.08 ^d^ |  |  | <0.001 | 0.09^d^ |  |  | 0.04 | 0.11^d^ |  |  | 0.58 | 0.10^d^ |
| <50,000 | 1260 (81.4) | 288 (18.6) |  |  | 915 (79.4) | 238 (20.6) |  |  | 283 (87.6) | 40 (12.4) |  |  | 62 (86.1) | 10 (13.9) |  |  |
| 50,000 – 100.000 | 946 (82.5) | 201 (17.5) |  |  | 712 (83.4) | 142 (16.6) |  |  | 175 (80.5) | 42 (19.4) |  |  | 59 (77.6) | 17 (22.4) |  |  |
| 100.000 – 150,000 | 466 (78.3) | 129 (21.7) |  |  | 363 (78.6) | 99 (21.4) |  |  | 70 (76.1) | 22 (23.9) |  |  | 33 (80.5) | 8 (19.5) |  |  |
| >150,000 | 489 (87.9) | 67 (12.1) |  |  | 412 (88.8) | 52 (11.2) |  |  | 42 (80.8) | 10 (19.2) |  |  | 35 (87.5) | 5 (12.5) |  |  |
| Prefer not to say | 652 (78.1) | 183 (21.9) |  |  | 482 (77.1) | 143 (22.9) |  |  | 126 (79.7) | 32 (20.3) |  |  | 44 (84.6) | 8 (15.4) |  |  |
| **English as primary language** |  |  | <0.001 | 0.07 |  |  | <0.001 | 0.11 |  |  | 0.24 | -0.04 |  |  | 0.43 | 0.06 |
| Yes | 3437 (82.4) | 735 (17.6) |  |  | 2668 (82.4) | 570 (17.6) |  |  | 554 (81.8) | 123 (18.2) |  |  | 215 (83.7) | 42 (16.3) |  |  |
| No | 376 (74.0) | 132 (26.0) |  |  | 216 (67.7) | 103 (32.3) |  |  | 142 (86.1) | 23 (13.9) |  |  | 18 (75.0) | 6 (25.0) |  |  |
| **Location** |  |  | <0.001 | 0.06 |  |  | <0.001 | 0.08 |  |  | 0.60 | -0.02 |  |  | 0.14 | 0.10 |
| Metropolitan location | 2686 (83.1) | 548 (16.9) |  |  | 1987 (83.1) | 403 (16.9) |  |  | 497 (82.1) | 108 (17.9) |  |  | 202 (84.5) | 37 (15.5) |  |  |
| Regional/Rural location | 1127 (77.9) | 320 (22.1) |  |  | 897 (76.8) | 271 (23.2) |  |  | 199 (84.0) | 38 (16.0) |  |  | 31 (73.8) | 11 (26.2) |  |  |
| ^a^ P-value for Chi-square differences by vaccination status. Gender did not include non-binary/other categories and highest educational level did not include other category.  ^b^ Phi coefficient.  ^c^ Independent samples t-tests and eta squared were used to measure differences between vaccination status and effect size for age as a continuous variable, respectively.  ^d^ Cramer’s V reported.  Significant differences between vaccinated and not vaccinated for patients who identify as Aboriginal and/or Torres Strait Islander were detected only for: 1) all diseases (p<0.001) and; 2) cancer (p=0.003). AUD = Australian Dollars. | | | | | | | | | | | | | | | | |

### Figure S1. Response frequencies for each DIVAS-6 item, by chronic disease type and vaccination status (a-f). ‘Don’t know’ responses have been excluded.

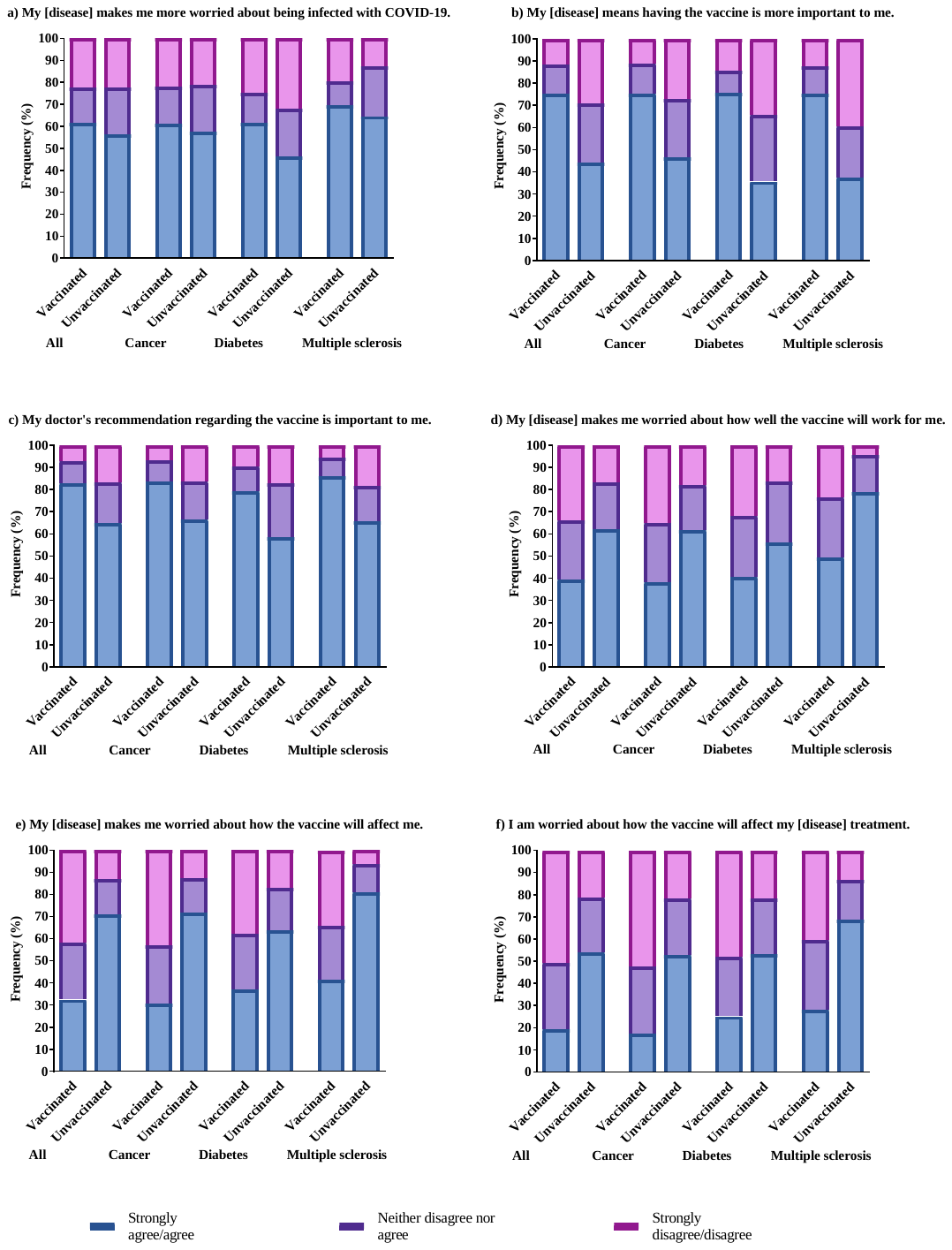
